# Standardising Breast Radiotherapy Structure Naming Conventions: A Machine Learning Approach

**DOI:** 10.1101/2022.10.14.22280859

**Authors:** Ali Haidar, Matthew Field, Vikneswary Batumalai, Kirrily Cloak, Daniel Al Mouiee, Phillip Chlap, Xiaoshui Huang, Vicky Chin, Farhannah Aly, Martin Carolan, Jonathan Sykes, Shalini K. Vinod, Geoffrey P Delaney, Lois Holloway

## Abstract

In progressing the use of big data in health systems, standardised nomenclature is required to enable data pooling and analyses. In many radiotherapy planning systems and their data archives, target volumes (TV) and organ-at-risk (OAR) structure nomenclature has not been standardised. Machine learning (ML) have been utilized to standardise volumes nomenclature in retrospective datasets. However, only subsets of the structures have been targeted. Within this paper, we proposed a new approach for standardizing all the structures nomenclature by using multi-modal artificial neural networks. A cohort consisting of 1613 breast cancer patients treated with radiotherapy was identified from Liverpool & Macarthur Cancer Therapy Centres, NSW, Australia. Four types of volume characteristics were generated to represent each target and OAR volume: textual features, geometric features, dosimetry features, and imaging data. Five datasets were created from the original cohort, the first four represented different subsets of volumes and the last one represented the whole list of volumes. For each dataset, 15 sets of combinations of features were generated to investigate the effect of using different characteristics on the standardisation performance. The best model reported 99.416% classification accuracy over the hold-out sample when used to standardise all the nomenclatures in a breast cancer radiotherapy plan into 21 classes. Our results showed that ML based automation methods can be used for standardising naming conventions in a radiotherapy plan taking into consideration the inclusion of multiple modalities to better represent each volume.

## I. Introduction

**R**ADIOTHERAPY data are used in various clinical research questions aiming to improve patients’ treatment and assess patterns of care such as dosimetry analyses, outcome modelling, toxicity, and automated contouring [1-5]. Radiotherapy data are large and require extensive amounts of time to clean and process. According to Dasu and Johnson, 80% of the time in data analytical research is taken up by data cleaning, curation, and preparation of medical records [6].

In breast cancer radiotherapy, individualised treatment plans are developed to optimise each patient’s radiation dose delivery. The patient’s restricted-dose organs-at-risk (OAR) and high-dose tumour target volumes (TV) are defined, together with additional regions-of-interest (ROIs) belonging to other categories such as machine specific ROIs, optimization structures, and control structures. OARs include the heart, left lung, right lung, combined lung, and the contralateral breast. TVs include the breast clinical target volume (CTV) and planning target volume (PTV), chest wall CTV and PTV, nodal CTVs and PTVs. Control structures include planning risk volumes (PRV) (e.g. heart PRV, lung PRV). Other contours include various number of ROIs (e.g. 2_Elekta_Shell_0, external, RING).

As shown in Fig. 1, inconsistency has been observed in the OAR and TV nomenclature in retrospective datasets. Standardised approaches are required to classifying TV and OARs in any cancer site, to utilise big data for radiotherapy applications and enable data pooling and analyses. In many radiotherapy planning systems and retrospective datasets, OAR and TV nomenclature has not been standardised. This inconsistency might be due to several reasons, such as the lack of specific templates or protocols for structure naming, variability in naming conventions between institutions, clinicians’ lack of adherence to naming protocols and spelling errors. Furthermore, nomenclature may change with time as new radiotherapy techniques are implemented. Despite strict protocols, inconsistency in structure names have been also recognized as an issue in clinical trials [7].

**Fig. 1.**
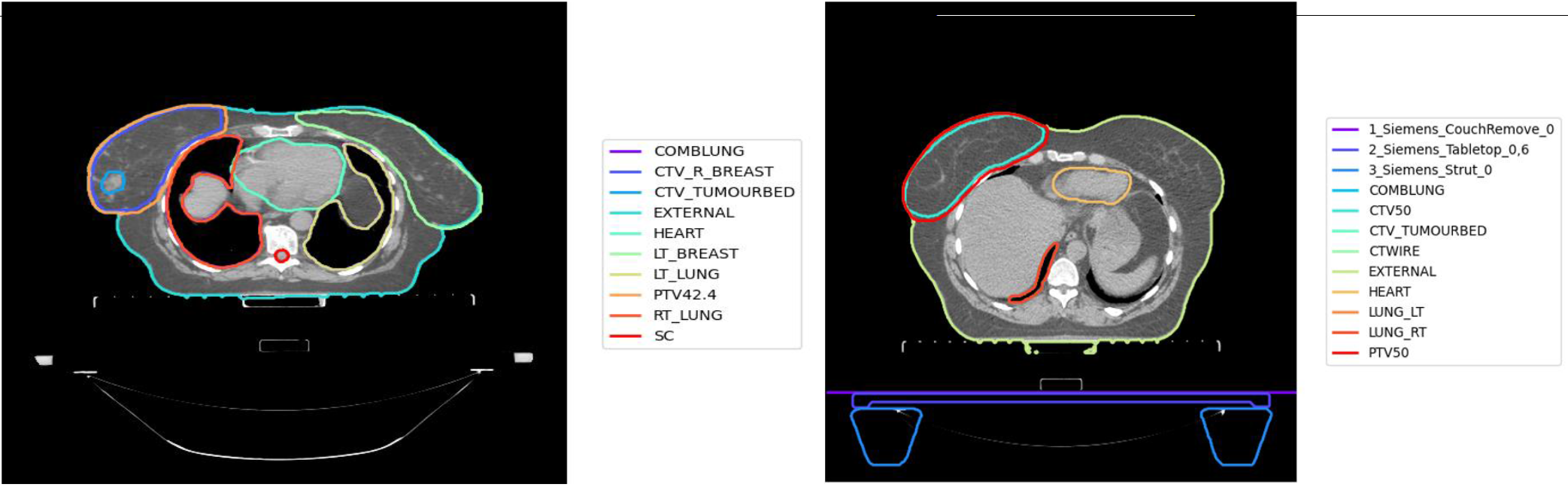
Variations in some OARs and TVs for two breast cancer patients. Left image: breast ctv named CTV_R_BREAST; left lung named LT_LUNG; right lung named RT_Lung. Right image: breast ctv as CTV_50; left lung as LUNG_LT; right lung as LUNG_RT.

To address this issue, health institutions released protocols for standardizing treatment structures at certain timeframes [8, 9]. To handle inconsistency in naming structures, the American Association of Physicists in Medicine (AAPM) developed a protocol for standardizing structure names to enable data pooling in multiple areas such as outcomes research, registries, and clinical trials, known as TG-263 [10]. However, patients treated before the release of these protocols still require standardization of OARs and TVs for inclusion in retrospective studies. Furthermore, some limitations in treatment planning software character limits the way structure names are displayed during the treatment planning process and has resulted in these protocols being incompletely implemented. There are also challenges for centres not using English for structure naming.

One possible method to handle inconsistency in naming structures is to manually check each patient plan and relabel the structures into standardized names. This is considered expensive and unachievable, in platforms that utilize large datasets from multiple institutions, such as The Australian Computer Aided Theragnostics network (AusCAT), which is a framework that was established across national and international radiation oncology departments, to enable data mining and learning from clinical practice datasets [11].

Another possible solution is to develop rule-based systems following discussion with clinicians. To handle variations in each environment, clinical staff and researchers are usually involved. The process includes discussion between the clinical staff and the researchers responsible for preparing the datasets to write scripts to interpret the variations. This entails time/effort from each of the involved parties. The variations are then grouped under a standardized name for each structure. With new patients’ records added, the current rules are revisited and updated to match variations in the updated records. Hereafter, these rules gain complexity with time. Manual intervention is still required to validate the results, as no rules can cover the whole set of conditions. This process is also considered expensive and time consuming, especially with datasets from multiple institutions. Considering the time and effort required for standardizing nomenclature, we observed the need for new systems to automate the process of standardizing OARs and TVs naming.

In recent years, machine learning (ML) algorithms have been incorporated to standardize OARs and TVs nomenclature over multiple types of cancer patients’ datasets. This included lung, prostate and head and neck cancer datasets [9, 12, 13], but to our knowledge, no model has yet been developed to standardise nomenclature in breast cancer radiotherapy data. These methods utilized various types of ML algorithms such as convolutional neural networks (CNNs), gradient-boost machine (GBM), and multi-layered perceptron (MLP). In these studies, 2D images, 3D volumes, extracted features from volumes and images, or extracted features for text were used as input to the models trained to standardise naming conventions. The developed studies did not consider all the TVs and OAR used in treatment plan, which is needed for real world applications. Hereafter, we propose a new approach for standardising nomenclature that can be utilised throughout the whole list of breast radiotherapy plan volumes, as well as portions of it by using artificial neural networks (ANNs). Section II reviews related work; Section III describes materials and methods; Section IV reports experiments and results; Section V derives a conclusion.

## II. Related Work

Rozario et al. conducted a feasibility study, which was among the first studies that incorporated ML algorithms to automate standardizing nomenclature, using a CNN to standardize organ labelling in prostate and head and neck cancer datasets [12]. Five OARs were used in the prostate dataset and nine OARs in the head and neck cancer dataset. 2D images were extracted and used as input to the CNN. 100% classification accuracy was reported with the proposed approach, but no TV have been considered. Hence, TV and other structures should be handled before running the developed model.

Another framework was proposed to standardize OARs [14, 15], which utilized an ensemble of CNNs in head and neck cancer patients (3DNNV). The framework consisted of multiple ResNets, which is a CNN originally trained on the ImageNet dataset, with non-local blocks that were combined using majority voting [16]. The authors proposed adaptive sampling and adaptive cropping (ASAC) to scale and crop the images as input to the networks. Three cohorts were utilized in the study, one for training and the rest for testing. 28 head and neck OARs were selected for modelling in this study. 3D volumes were used as input to the ensemble components. Several baseline models were introduced for comparison and analyses purposes. These models were compared to the proposed framework, which reported better performance in terms of three evaluation metrics: true positive rates (TPR), area under the curve (AUC) of the receiver operating characteristics (ROC), and f1-score. The proposed framework was also compared to two alternative approaches, the first uses a fuzzy string-matching algorithm while the other uses a five layered CNN. The authors trained the five layered CNN using their dataset. However, the fuzzy model was used for testing only. Better performance was obtained with the proposed framework compared to the two alternative approaches. However, these studies lack the use of TV such as the Gross-Tumour Volume (GTV), PTV, and CTV.

Text features have also been used in standardising nomenclatures. Syed et al. utilized ML algorithms to standardize OARs in lung and prostate cancer datasets into TG-263 standardised names [17]. A dataset that consisted of 794 prostate and 754 lung cancer patients from 40 different centres managed by the Veterans health administration (VA) was used for developing the model. Another dataset was collected from the radiation oncology department at Virginia Commonwealth University (VCU) and used as the hold-out sample. 10 prostate and 9 lung OAR were identified in the study. The other structures were named as non_OAR. The structure names were processed and manipulated to numerical representations, then an algorithm named fastTEXT was used for training the collected records. The authors reported an f1-score of 0.93 for prostate structures and 0.95 on lung structures over the hold-out samples. Similarly, the study did not consider TV, which usually adds the highest levels of complexity in the standardization process.

Sleeman et al. proposed an approach to standardize structures based on volumetric bitmap representations and five different ML algorithms [9]. Prostate and lung cancer datasets were included in the study. Apache spark was used to train the multi-centred datasets across 40 different institutions. A dataset that consisted of 1200 patients was used for training and validation, while a dataset that consisted of 100 patients was used for testing (50 lung, 50 prostate). Five structures were annotated for the lung patients and seven for the prostate patients. Two datasets were created: curated dataset which only contains the selected structures, and non-curated data which contains everything expected in a study. Bitmap images were created and converted to feature vectors. Two types of images were created with each dataset (curated/non-curated): one contained the patient’s bone anatomy, and the other did not. The generated feature vectors consisted of hundreds of thousands of features, which required the inclusion of dimensionality reduction through truncated singular value decomposition (SVD). The records were reduced to 100 input features. Five different classifiers were used in this study: naïve bayes (NB), random forest (RF), gradient-boost machine (GBM), multi-layered perceptron (MLP), and support vector machine (SVM). The datasets were manipulated to obtain balanced samples for each of the included organs. The f1-score was used to measure the performance of each classifier. The MLP outperformed the other algorithms in majority of the tests over the curated datasets. The results showed improved results with the inclusion of bone anatomy in the datasets. Only one target volume (PTV) was utilized in this study. The highest accuracy achieved by a model was less than 92% when using the non-curated datasets.

## III. Materials and methods

### A. Data Collection and Labelling

A dataset consisting of 1613 left/right breast cancer patients treated between 2014 and 2018 was collected from Liverpool & Macarthur Cancer Therapy Centres, New South Wales, Australia. This study was approved by the NSW Population & Health Services Research Ethics Committee (2019/ETH01550; 11/09/2016). Each patient’s radiotherapy treatment plan consisted of a set of volumes with inconsistent names over the whole cohort. Several discussions with the clinicians at the centres were required to label the cohort. The labels were clustered into five different groups shown in TABLE I.

**TABLE I.** standardised labels and categories of the structures.

| Nb | Class | Category | Unique values |
| --- | --- | --- | --- |
| 1 | Combined lungs | OAR | 17 |
| 2 | Contralateral breast | OAR | 21 |
| 3 | Heart | OAR | 3* |
| 4 | Left lung | OAR | 9 |
| 5 | Right lung | OAR | 9 |
| 6 | Breast CTV | TV | 89 |
| 7 | Breast PTV | TV | 161 |
| 8 | Chestwall CTV | TV | 19 |
| 9 | Chestwall PTV | TV | 13 |
| 10 | Tumourbed CTV | TV | 30 |
| 11 | Tumourbed PTV | TV | 12 |
| 12 | Axilla CTV | Nodal TV | 42 |
| 13 | Axilla PTV | Nodal TV | 4 |
| 14 | Imc CTV | Nodal TV | 18 |
| 15 | Imc PTV | Nodal TV | 16 |
| 16 | Scf CTV | Nodal TV | 23 |
| 17 | Scf PTV | Nodal TV | 17 |
| 18 | Control structure | Control | 120 |
| 19 | Boost structure | Boost | 36 |
| 20 | Combined structure | Comb TV | 16 |
| 21 | Exclude | exclude | 328 |

\*low number of variations was observed with the naming conventions of the heart volumes.

### B. Input Features Generation

Discussions with clinicians about how they determined what a non-conventionally labelled structure was led to several observations. Initially, the clinicians tend to look at the text to categorize each structure. The structure image might be checked and visualized for further interpretation. Furthermore, the position of the structure may be analysed. Finally, the dosimetry values might be checked for analysing the structure name. We aimed to utilize various types of features to mimic the approach followed by the clinicians with the use of neural networks that are originally inspired by the human biological brain. Four types of characteristics were generated for each structure/volume in each patient radiotherapy plan:

- **Textual features**: A treatment plan consists of volumes with associated names. These volumes/structures are defined using alphanumerical characters that represent various patient structures. A set of features was created by highlighting the existence of specific text blocks in each structure. e.g. a feature ‘breast’ was introduced where a value of 1 was associated with all the structures that contained the text ‘breast’, otherwise a value of 0 was allocated. 15 features were introduced to represent the occurrence of substrings in structure names (breast, lung, chest wall or cw, axilla or ax, etc.). Five additional features were created that summarises: number of letters in a structure name, number of digits in a structure name, number of spaces in a structure name, number of other characters in a structure name (commas, underscores, etc.), and the total number of characters in a structure name. In total, 20 textual features were extracted from each structure/volume text.
- **Imaging features:** 2D central slices were created to include details about the structure shape, size, and imaging biomarkers in modelling. The central slice was defined as the CT slice with the highest number of tumour pixels on the z-axis. Each structure binary mask was overlaid on the CT image to select the pixel values of the central slice. The Hounsfield Units (HU) representing pixel values inside the structure were selected, while pixel values surrounding the structure were replaced by zeros.
- **Geometry features**: Positional features were extracted to include details of the position of each volume. The coordinates of the centroid of the 3D volume were calculated and included as features. For each structure, the magnitude of the vectors connecting the centroid of the structure to the point (0, 0, 0) in the three-dimensional space were calculated. The directions over each axis (cosine angles) of the magnitude vector were also calculated. The index of the central slice over the z-axis was also included. Finally, the number of voxels representing the structure was included. In total, nine positional and volumetric features were used: x, y, and z coordinates of the centroid centroid; cosine angles on each dimension of the magnitude vector connecting (0, 0, 0) to the centroid; magnitude; number of voxels; index).
- **Dosimetry features**: A total of 10 dosimetry features were calculated for each structure: minimum dose, median dose, mean dose, maximum dose, V%20.0,V%10.0,V%5.0,V%95.0,V%105.0,V%110.0, D50.0, where Vx is the dose received by x% of the volume and Dy is the volume receiving at least y% of the prescription dose.

### C. Datasets Generation and Pre-processing

Five datasets were created from the original cohort, the first four represented different subsets of volumes and the last one represented the whole list of volumes. The categories of the classes used in each study are summarised in TABLE II (further details in Appendix A). The first dataset represents a case study where OARs were targeted for standardisation. In other words, if methods have already been prepared to standardise all/some of the other categories (TV, nodes TV), and there is no need to utilize a full model. With Dataset5, it was assumed that no subsets have been standardised and there is a need to identify any possible volume in the patient radiotherapy plan.

**TABLE II.** Datasets

| Dataset | category | nb of classes | classes |
| --- | --- | --- | --- |
| Dataset 1 | OARs | 5 | heart, combined lungs, left lung, right lung, contralateral breast |
| Dataset 2 | OARs + TVs | 11 | heart, combined lungs, left lung, right lung, contralateral breast, breast CTV, breast PTV, chestwall CTV, chestwall PTV, tumourbed PTV, tumourbed CTV |
| Dataset 3 | OARs + TVs + nodes TVs | 17 | heart, combined lungs, left lung, right lung, contralateral breast, breast CTV, breast PTV, chestwall CTV, chestwall PTV, tumourbed CTV, tumourbed PTV, scf CTV, scf PTV, axilla PTV, axilla CTV, imc CTV, imc PTV |
| Dataset 4 | OARs+ TVs+ nodes TVs + combined structure + boost structures + control structures | 20 | heart, combined lungs, left lung, right lung, contralateral breast, breast CTV, breast PTV, chestwall CTV, chestwall PTV, tumourbed CTV, tumourbed PTV, scf CTV, scf PTV, axilla PTV, axilla CTV, imc CTV, imc PTV, combined structure, control structure, boost structure |
| Dataset 5 | OARs+ TVs+ nodes TVs + combined structure + boost structures + control structures + excluded | 21 | heart, combined lungs, left lung, right lung, contralateral breast, breast CTV, breast PTV, chestwall CTV, chestwall PTV, tumourbed CTV, tumourbed PTV, scf CTV, scf PTV, axilla PTV, axilla CTV, imc CTV, imc PTV, combined structure, control structure, boost structure, exclude |

To build ML models, the datasets were partitioned into training, validation, and test samples. The training dataset was used to train the algorithm. The validation dataset was used to track the model performance and to avoid overfitting. The test dataset was used to evaluate the performance of the developed model on unseen samples.

The original dataset consisted of 1613 patients. 173 patients were selected as the test dataset, while 1440 patients were used for training and validating the ML algorithms. The number of classes varied per patient. We targeted the selection of a stratified cohort, where at least 10% of each class will be obtained in the test cohort in each study, e.g. the total number of patients with internal mammary lymph node (imc) PTV volumes was five, one sample was guaranteed for testing. In addition, for all the datasets, the same 173 patients were used for evaluation.

ANNs are known to perform better with smaller ranges. Three types of features were tabular and one was image data (central slices). Text, dose, and geometrical features were normalized into smaller ranges (between 0 and 1). As mentioned earlier, the pixel values in the imaging data were described in HU. For image data, lower (−255) and upper (+255) HU bounds were applied over each pixel value in the central slices. The central slices were then resized into 64*64 before being mapped into values between 0 and 1. Values surrounding the structure were reserved as 0 without being altered in the normalisation process.

While labelling the patients datasets, it was noticed that a class might occur one or more times in each plan, e.g. a patient might have a structure named ctv42,4 and another structure named ctv_l_breast. The two structures will be interpreted as left_breast_ctv, however, one will be selected as final. In real case scenarios, this kind of situation is expected. For this reason, we removed the duplicates/alternatives from the training dataset (to explicitly train the model to detect similar patterns in new datasets), but for evaluation purposes we introduced two datasets:

- Original test dataset: consists of the final selected structures by the clinicians
- Extended test dataset: consisted of all the structures in the dataset, with the final being flagged.

### D. Artificial Neural Networks (ANNs)

An ANN is a ML algorithm inspired by the human biological brain. It processes information through multiple processing units, known as neurons, distributed over multiple layers to explore patterns and trends in data. To learn from data, the ANN is trained for a number of rounds, known as epochs, where the data samples are shown repeatedly to the ANN architecture in an attempt to minimise the error between the actual and predicted output. Neural networks are trained by updating weights and bias connecting the information processing units (neurons) across layers. Deep learning is a branch in ML where information are processed in a neural network through multiple layers (4 or more) of neural processing units.

The ANN can be adapted to accept any type of input data such as tabular, images, and multi-modal records. In neural networks where the input data consists of numerical features, fully connected layers are typically utilized to form a Feed Forward Neural Networks (FFNN). In neural networks where the input data consists of images, blocks of convolutional and pooling layers are utilized to form a CNN. With multi-modal input, both fully connected layers and convolutional blocks are used to form a Multi-input neural networks (MINN).

For each of the five datasets, a total of 15 experiments were conducted representing the combinations of the four types of input features. FFNNs were utilised with seven case studies that used tabular data (Fig. 1a): text, dose, geometry, text + dose, text + geometry, dose + geometry, text + dose + geometry. A CNN was used with the case study that utilised images only (Fig. 2b). To integrate the textual, geometrical, dosimetric, and imaging data, multi-input deep ANNs were utilized (Fig. 2c). MINNs were utilised with seven case studies that used tabular and imaging data: text + image, dose + image, geometry + image, text + dose + images, text + geometry + images, dose + geometry + images, text + dose + geometry +images.

**Fig. 2.**
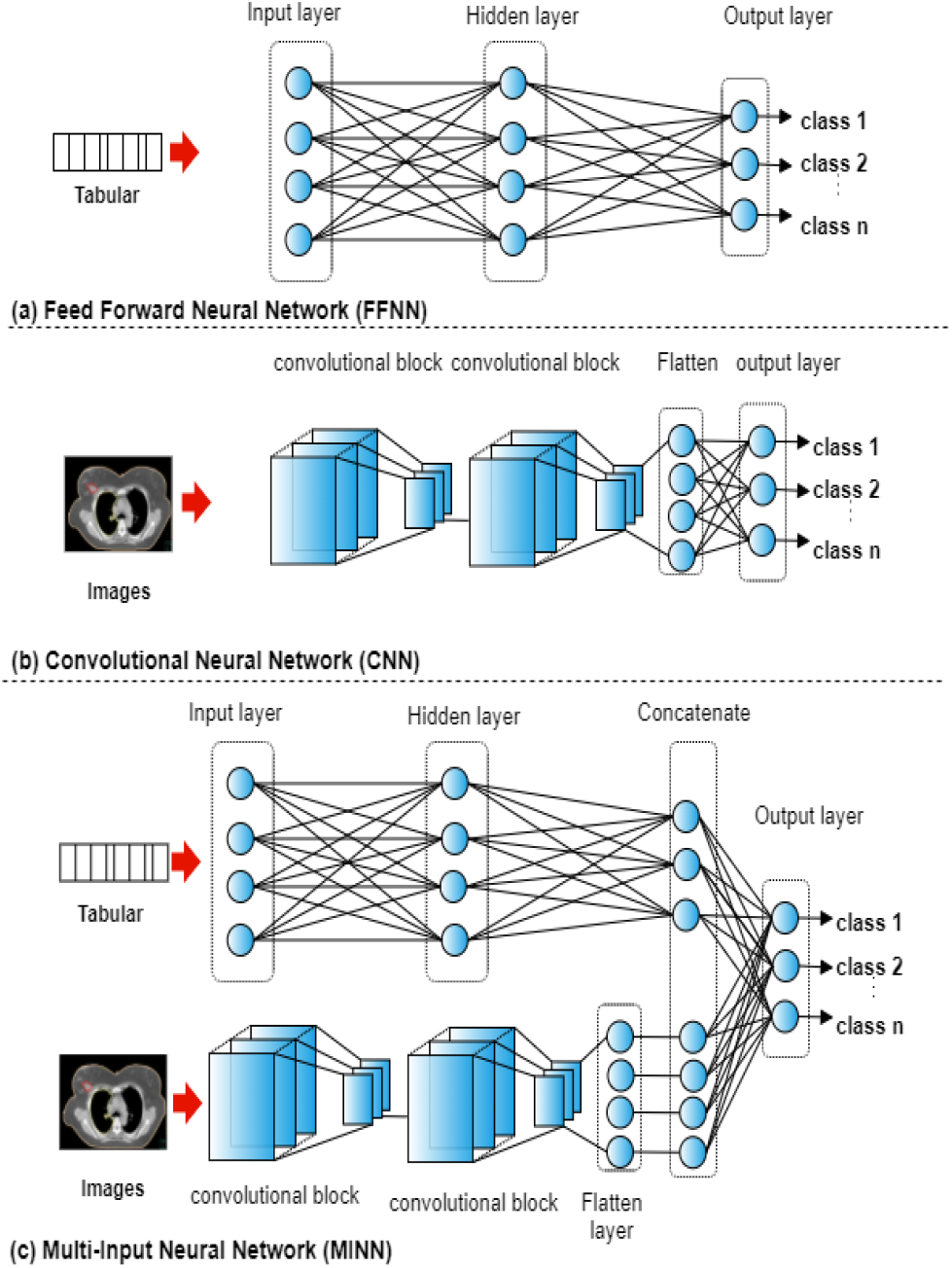
An overview of the utilized neural networks.

The network architectures were the same across each of the case studies. The FFNN consisted of three layers with 18 neurons in the hidden layer and sigmoid as the activation function. The CNN consisted of two convolutional blocks (convolutional, pooling, and dropout layers [18]) followed by a flatten layer and an output layer as shown in Fig. 2c. The MINN combined the FFNN and CNN architectures by removing the last layer, adding a concatenation layer followed by an output layer. The ‘softmax’ activation function as used in the output layer with the three architectures. In neural networks with mixed input data, a model f is trained by utilizing input features that belong to different categories. The output p of the network is defined as:

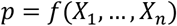

Where *X*_*i*_ in an input modality, 1 < *i* ≤ *n* n is the number of input modalities the deep network can receive. The training parameters for each network are shown in Table III.

**TABLE III.** Parameters for training the ANNS.

| parameter | value |
| --- | --- |
| Epochs | 120 |
| Batch size | 16 |
| Optimizer | Adam |
| Learning rate | 0.01 |
| Early stopping | If 12 epochs with no improvement |
| Reduce learning rate | *0.1 if no improvement in 6 epochs |
| Loss function | Categorical cross-entropy |

## IV. Experiments and Results

### A. Experimental Setup

Four types of volumes characteristics were generated for each structure in each dataset (text, dose, geometry, and images). A total of 15 combinations of input features were generated, each representing an experiment in each dataset. Further details are shown in Appendix B. A windows server with 10 virtual CPUs and 40GB of RAM was utilized to prepare the datasets and to train the ANNs. Keras and Tensorflow were used for developing the neural networks [19, 20].

### B. Results and Analyses

Classification accuracy over the original and extended test datasets of each developed model in each dataset for each combination of features are shown in TABLE IV and TABLE V. Similar to other literature, standardising OAR only (Dataset1) can be achieved using only imaging data. Any model with images as input showed a 100% accuracy when modelling OARs only. Adding the TV (Dataset 2 & 3) highlighted the need to include more than one type of features for standardising TV nomenclatures. With dose and positional features, the feature characteristics for structures (PTVs & CTVs) are quite similar and would require additional features for identification. It was noticed that using text and images is mandatory to achieve reliable models. As expected, combining multiple features revealed higher classification accuracy compared to using single features. Reliable performance was observed with all the datasets when using the text feature as input to the model, which aligns with the traditional approach, where clinicians tend to look at text initially to standardise nomenclatures.

**TABLE IV.**
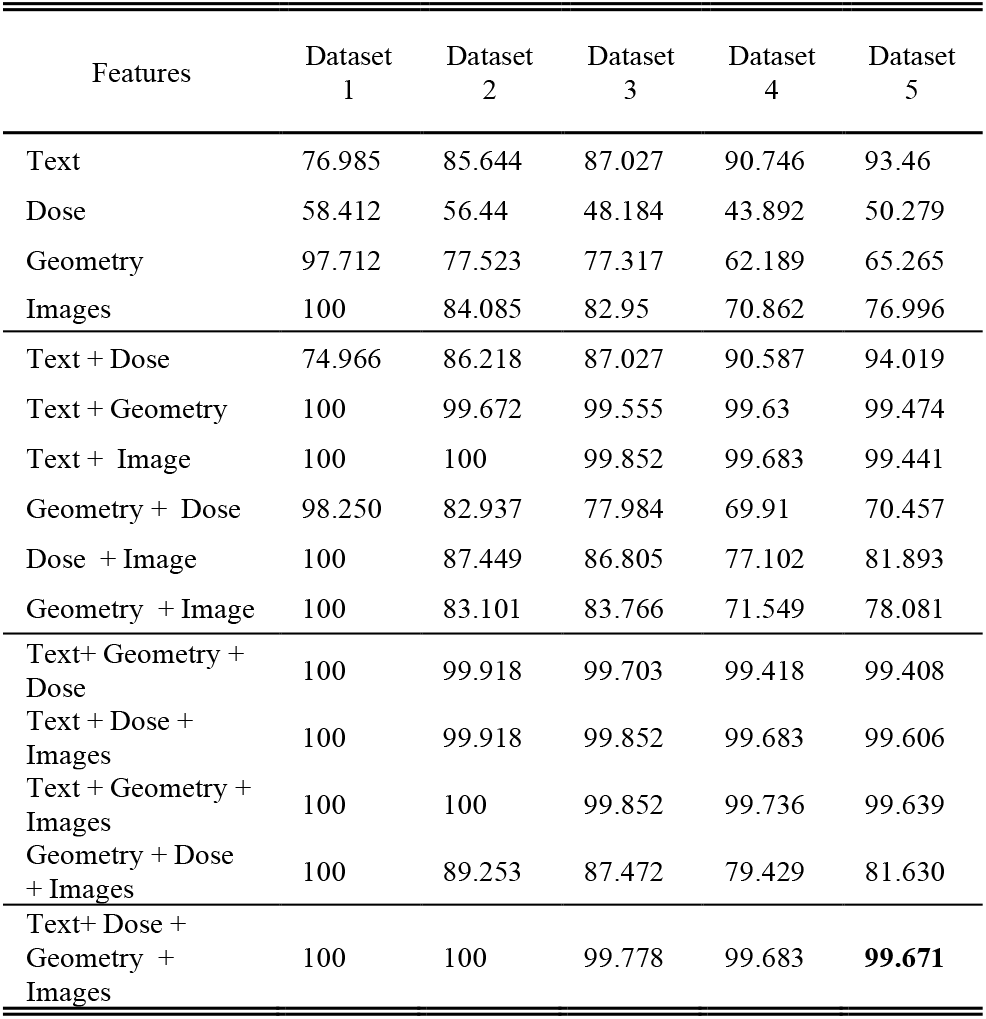
Classification accuracy over the test dataset.

| Features | Dataset 1 | Dataset 2 | Dataset 3 | Dataset 4 | Dataset 5 |
| --- | --- | --- | --- | --- | --- |
| Text | 76.985 | 85.644 | 87.027 | 90.746 | 93.46 |
| Dose | 58.412 | 56.44 | 48.184 | 43.892 | 50.279 |
| Geometry | 97.712 | 77.523 | 77.317 | 62.189 | 65.265 |
| Images | 100 | 84.085 | 82.95 | 70.862 | 76.996 |
| Text + Dose | 74.966 | 86.218 | 87.027 | 90.587 | 94.019 |
| Text + Geometry | 100 | 99.672 | 99.555 | 99.63 | 99.474 |
| Text + Image | 100 | 100 | 99.852 | 99.683 | 99.441 |
| Geometry + Dose | 98.250 | 82.937 | 77.984 | 69.91 | 70.457 |
| Dose + Image | 100 | 87.449 | 86.805 | 77.102 | 81.893 |
| Geometry + Image | 100 | 83.101 | 83.766 | 71.549 | 78.081 |
| Text+ Geometry + Dose | 100 | 99.918 | 99.703 | 99.418 | 99.408 |
| Text + Dose + Images | 100 | 99.918 | 99.852 | 99.683 | 99.606 |
| Text + Geometry + Images | 100 | 100 | 99.852 | 99.736 | 99.639 |
| Geometry + Dose + Images | 100 | 89.253 | 87.472 | 79.429 | 81.630 |
| Text+ Dose + Geometry + Images | 100 | 100 | 99.778 | 99.683 | **99.671** |

**TABLE V.**
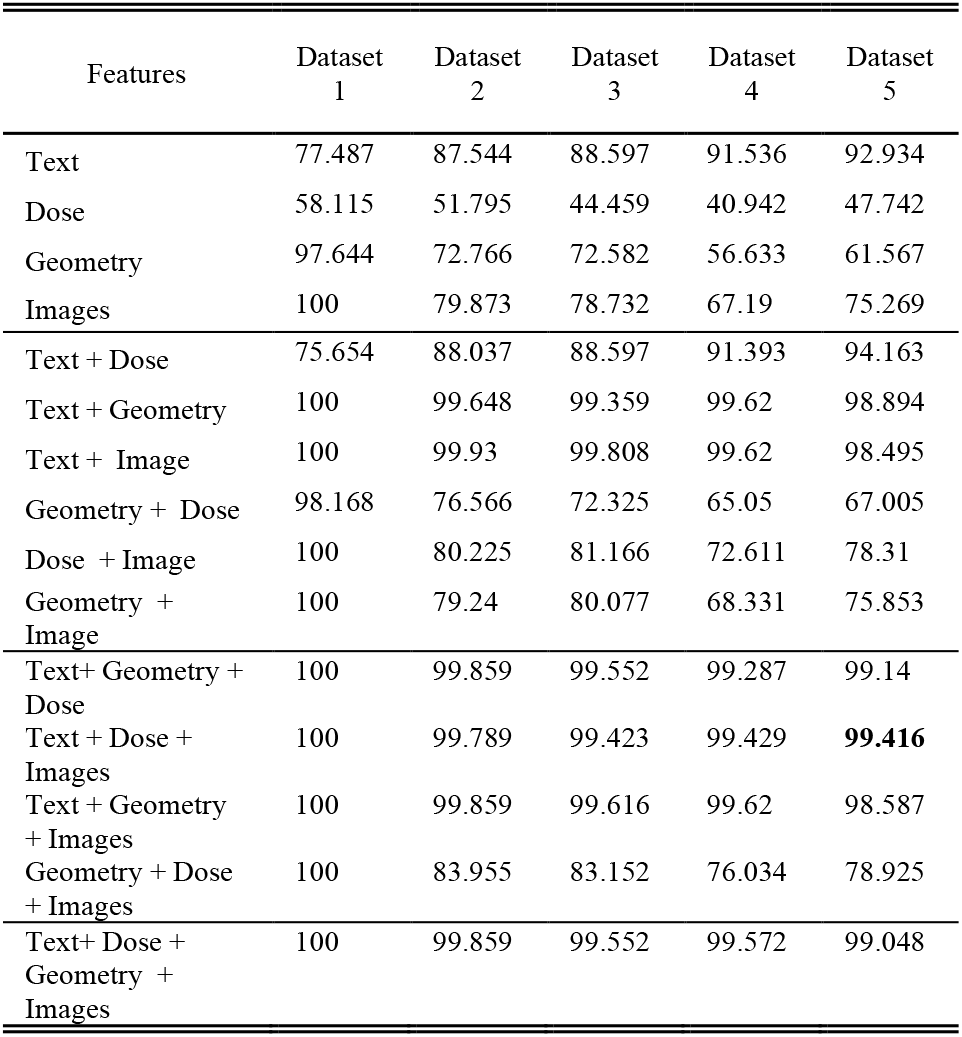
Classification accuracy over the extended test dataset.

| Features | Dataset 1 | Dataset 2 | Dataset 3 | Dataset 4 | Dataset 5 |
| --- | --- | --- | --- | --- | --- |
| Text | 77.487 | 87.544 | 88.597 | 91.536 | 92.934 |
| Dose | 58.115 | 51.795 | 44.459 | 40.942 | 47.742 |
| Geometry | 97.644 | 72.766 | 72.582 | 56.633 | 61.567 |
| Images | 100 | 79.873 | 78.732 | 67.19 | 75.269 |
| Text + Dose | 75.654 | 88.037 | 88.597 | 91.393 | 94.163 |
| Text + Geometry | 100 | 99.648 | 99.359 | 99.62 | 98.894 |
| Text + Image | 100 | 99.93 | 99.808 | 99.62 | 98.495 |
| Geometry + Dose | 98.168 | 76.566 | 72.325 | 65.05 | 67.005 |
| Dose + Image | 100 | 80.225 | 81.166 | 72.611 | 78.31 |
| Geometry + Image | 100 | 79.24 | 80.077 | 68.331 | 75.853 |
| Text+ Geometry + Dose | 100 | 99.859 | 99.552 | 99.287 | 99.14 |
| Text + Dose + Images | 100 | 99.789 | 99.423 | 99.429 | **99.416** |
| Text + Geometry + Images | 100 | 99.859 | 99.616 | 99.62 | 98.587 |
| Geometry + Dose + Images | 100 | 83.955 | 83.152 | 76.034 | 78.925 |
| Text+ Dose + Geometry + Images | 100 | 99.859 | 99.552 | 99.572 | 99.048 |

The confusion matrix represents the comparison between the predicted (x-axis) and true classes (y-axis). The confusion matrix for the best performing model with 99.671% classification accuracy over the original test sample is shown in Fig.3, where 0.329% (10 samples) of the volumes were misclassified by the developed model. 6 out of the 10 images were predicted not to use (i.e. Exclude) by the model. Further details are included Appendix B.

**Fig. 3.**
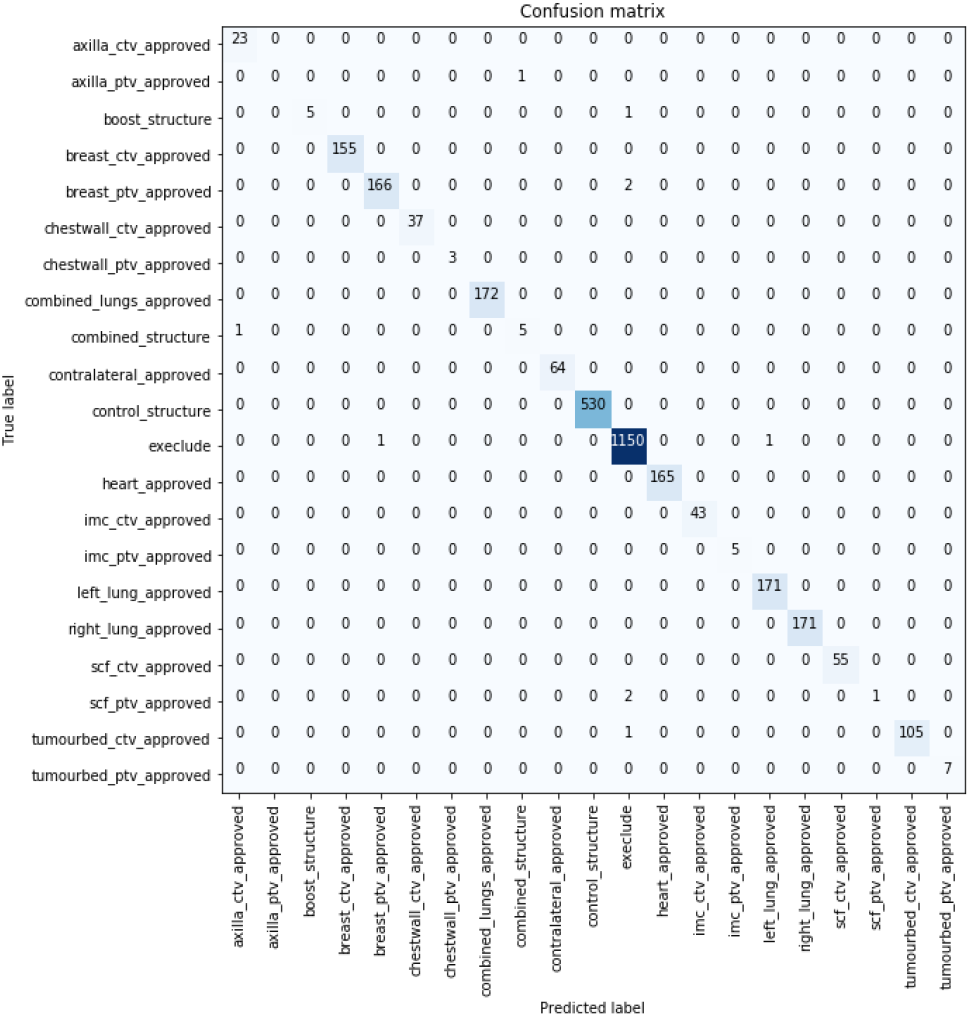
Confusion matrix of the best performing model over the test dataset

Similar performance was obtained with the models evaluated over the extended test datasets with the best performance being reported by model developed using (text + dose + images) with 99.416% classification accuracy. The confusion matrix for the best performing model over the extended dataset with 99.416% classification accuracy is shown in Fig.4. 19 samples were misclassified by this model, with more than half of them predicted as not to use by the model (i.e. exclude). Further details about the images are shown in Appendix B.

**Fig. 4.**
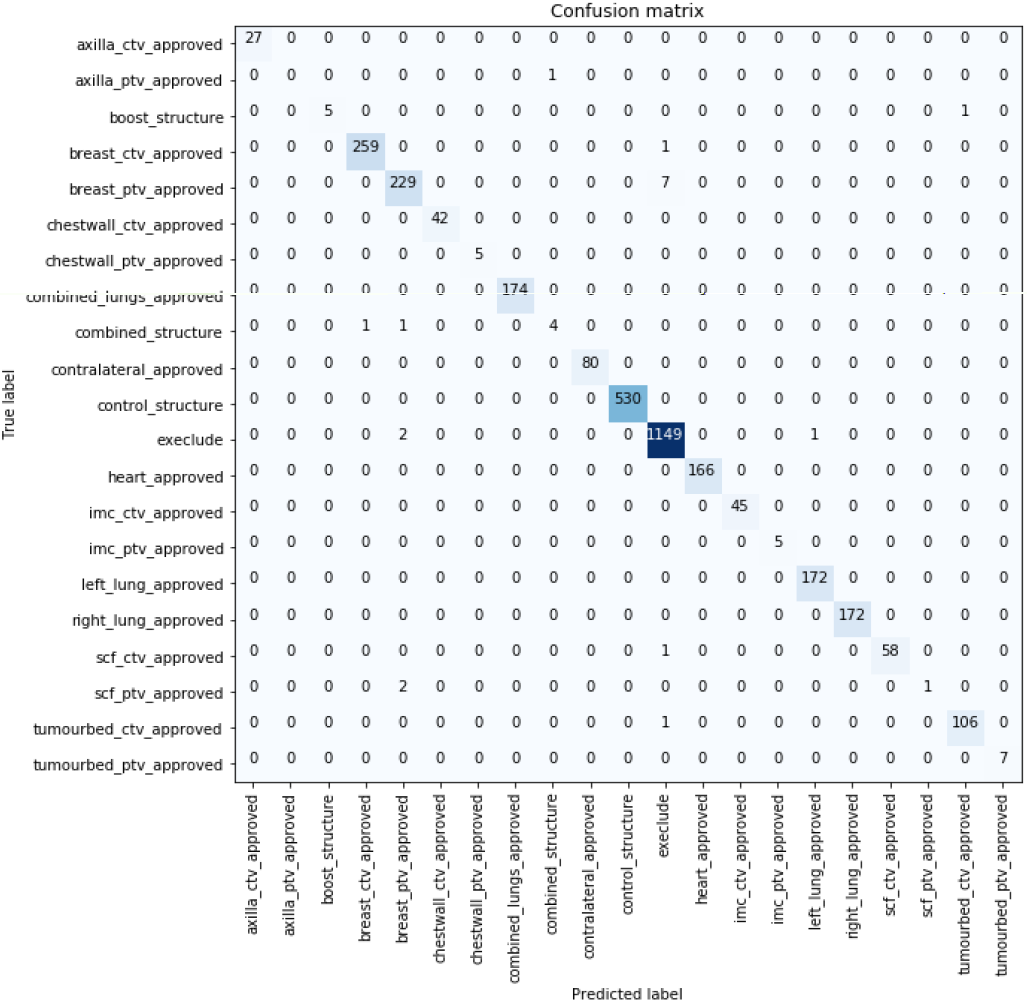
Confusion matrix of the best performing model extended test dataset.

The extended dataset might contain multiple structures in the same plan interpreted as the same TV/OAR. However, one structure will be used as the final structure for data mining analyses. While training, we selected only the structures that were reported as final by the clinicians. The other label referring to the same structure was assumed as a duplicate, registrar contour, etc. For the ‘breast PTV’, 40 patients in the extended test dataset had two or more volumes being referred as ‘breast PTV’ with one of them selected by the clinicians as final. We selected the structure with the highest probablity generated for each of the structures and referred to that as final. For 39 patients out of the 40, the structure selected as the final by the clinicians had the highest probablity in the patient plan. We assume that this approach worked because we trained the datasets using the final structures only.

As shown in Fig. 4 and 5, structures with lower numbers of occurences were misclassified, which highlights the need for more samples to be able to identify such structures.

### C. Discussion

We have developed different models for each dataset. With some datasets, the developed models might be used where some features are not possible to generate. With the high accuracy being reported with only two types of features, such models will be available for use in such cases.

In addition, there could be some volumes that have already been standardised such as the control structures. With the successful implementation of the models across the five types of datasets, such models can be used where there will be no need to standardise the already standardised volumes.

The time taken to train the two models that revealed the best performance over the two test datasets in Dataset 5 was less than 1.5 hours. Further details about the time taken to train each neural network for each study is shown in Appendix B. The training time was affected by the number of classes, convergence, early stopping, type of input features incorporated, and available hardware. The prediction time was in seconds.

It was noticed that the models converged quickly, which indicates that the extracted features of the volumes contains disciminative features. Within this study, we verified that supervised learning can be utilised in standardising breast cancer radiotherapy data. This showed potential of conducting the experiments using unsupervised learning. In addition, the datasets were used as is in this study with no undersampling on the list of structures.

Within this study, the test samples were collected from a single data centre. There is a need to further examine such models over additional datasets, which will be done as a part of our future work.

## V. conclusion

This paper presented an approach for standardising breast radiotherapy data by using artificial neural networks (ANNs). An original cohort consisting of 1613 patients was collected from Liverpool and Macarthur Cancer Therapy centres, NSW, Australia. Five datasets were created from the original cohort, representing different scenarios for standardizing radiotherapy volumes. Four types of features were extracted from each sample in the datasets: text, dosimetry, geometry features, and 2D images representing central slices. 15 different combinations of features were generated, and three types of neural networks were trained to standardize volumes. We conclude that the standardization of nomenclatures using ML is achievable taking into consideration the inclusion of multiple modalities while training the ML algorithm.

## Data Availability

All data produced in the present study are available upon reasonable request to the authors. Researchers will need to be added to the ethics protocol to access data used in this study.

### Appendices

- Appendix A: Standardised Breast Radiotherapy Structures Names
- Appendix B: Experiments and Results

## Acknowledgment

This work was supported by the South Western Sydney Local Health District (SWSLHD); Illawarra and Shoalhaven Local Health District (ISLHD); Western Sydney Local Health District (WSLHD); Nepean Blue Mountains Local Health District (NBMLHD); Ingham Institute for Applied Medical Research, Liverpool, NSW 2170, Australia; Ingham Institute Data and Cancer Research Grant 2019 “Detecting and Fixing Variations in Cancer Patients Medical Records”.

