## Appendices for "Standardising Breast Radiotherapy Structure Naming Conventions: A Machine Learning Approach"

### Appendix A. Standarised Breast Radiotherapy Structures Names

*Table A. 1 Classes utilized in each dataset.*

| Structure | Dataset 1 | Dataset 2 | Dataset 3 | Dataset 4 | Dataset 5 |
|---|---|---|---|---|---|
| axilla ctv | - | - | X | X | X |
| axilla ptv | - | - | X | X | X |
| boost structure | - | - | - | X | X |
| breast ctv | - | X | X | X | X |
| breast ptv | - | X | X | X | X |
| chestwall ctv | - | X | X | X | X |
| chestwall ptv | - | X | X | X | X |
| combined lungs | X | X | X | X | X |
| combined structure | - | - | - | X | X |
| contralateral | X | X | X | X | X |
| control structure | - | - | - | X | X |
| exclude | - | - | - | - | X |
| heart | X | X | X | X | X |
| imc ctv | - | - | X | X | X |
| imc ptv | - | - | X | X | X |
| left lung | X | X | X | X | X |
| right lung | X | X | X | X | X |
| scf ctv | - | - | X | X | X |
| scf ptv | - | - | X | X | X |
| tumourbed ctv | - | X | X | X | X |
| tumourbed ptv | - | X | X | X | X |
| Total | 5 | 11 | 17 | 20 | 21 |

### Appendix B. Experiments and Results

*Table B. 1 Experiments conducted in each case study.*

| Case study | Features | Type of the network |
|---|---|---|
| 1 | Text | Feed forward neural network |
| 2 | Dose | Feed forward neural network |
| 3 | Geometry | Feed forward neural network |
| 4 | Images | Convolutional neural network |
| 5 | Text + dose | Feed forward neural network |
| 6 | Text + geometry | Feed forward neural network |
| 7 | Text + images | Multi-input neural network |
| 8 | Geometry + dose | Feed forward neural network |
| 9 | Dose + image | Multi-input neural network |
| 10 | Geometry + image | Multi-input neural network |
| 11 | Text + geometry + dose | Feed forward neural network |
| 12 | Text + dose + image | Multi-input neural network |
| 13 | Text + geometry + image | Multi-input neural network |
| 14 | Geometry + dose +image | Multi-input neural network |
| 15 | Text + dose + geometry +image | Multi-input neural network |

*Table B. 2 list of misclassified volumes predicted by the best performing model over the test dataset (study 5).*

| Structure name | Structure category/class | Predicted category/class name | Probability |
|---|---|---|---|
| PTV_RTAXILLA | axilla_ptv_approved | combined_structure | 0.611028 |
| SCLAV_PTV | scf_ptv_approved | exclude | 0.704626 |
| SCLAV_PTV | scf_ptv_approved | exclude | 0.520875 |
| RO_SEROMA | tumourbed_ctv_approved | exclude | 1 |
| PTV42,4old | exclude | breast_ptv_approved | 0.975655 |
| ChestwallAx_CTV | combined_structure | axilla_ctv_approved | 0.81999 |
| lung0.5 | exclude | left_lung_approved | 0.999727 |
| PTV_NODES_02 | breast_ptv_approved | exclude | 0.717039 |
| L_BTREAST_PTV | breast_ptv_approved | exclude | 0.987708 |
| LEVEL_3_NODE_CTV_BOOST | boost_structure | exclude | 0.934735 |

*Table B. 3 list of misclassified samples by the best performing model over the extended test dataset.*

| Structure name | Structure category/class | Predicted category/class name | Probability |
|---|---|---|---|
| PTV_RTAXILLA | axilla_ptv_approved | combined_structure | 0.669003 |
| SCLAV_PTV | scf_ptv_approved | breast_ptv_approved | 0.889554 |
| PTV42,4 | breast_ptv_approved | exclude | 0.536556 |
| PTV_EVAL_STARS | breast_ptv | exclude | 0.984457 |
| SCLAV_PTV | scf_ptv_approved | breast_ptv_approved | 0.692799 |
| RO_SEROMA | tumourbed_ctv_approved | exclude | 1 |
| PTV42,4old | exclude | breast_ptv_approved | 0.981575 |
| PTV_TUMOURBEDBOOST | boost_structure | tumourbed_ctv_approved | 0.729915 |
| EXT-PTV | breast_ptv | exclude | 0.999907 |
| ChestwallAx_CTV | combined_structure | breast_ctv_approved | 0.554923 |
| ChestwallAx_PTV | combined_structure | breast_ptv_approved | 0.557953 |

| | | | |
|---|---|---|---|
| lung0.5 | exclude | left_lung_approved | 0.992461 |
| CTV_IP_NODES | breast_ctv | exclude | 0.999103 |
| PTV60_RING | exclude | breast_ptv_approved | 0.786016 |
| L_BTREAST_PTV | breast_ptv_approved | exclude | 0.952107 |
| PTV42,4_EVAL | breast_ptv | exclude | 0.510468 |
| PTVLD_EVAL | breast_ptv | exclude | 0.945179 |
| PTVLD_EVAL | breast_ptv | exclude | 0.892079 |
| REG_CTV_L_SCV | scf_ctv | exclude | 0.999954 |

*Table B. 4 Time taken to train each model in each dataset (in seconds).*

| case study | dataset 1 | dataset 2 | dataset 3 | dataset 4 | dataset 5 |
|---|---|---|---|---|---|
| text | 120 | 196 | 129 | 303 | 465 |
| dose | 125 | 191 | 204 | 293 | 479 |
| geometry | 124 | 183 | 242 | 301 | 478 |
| images | 933 | 2346 | 2481 | 3872 | 9782 |
| text_dose | 62 | 106 | 193 | 272 | 432 |
| text_geometry | 88 | 155 | 274 | 262 | 465 |
| text_image | 685 | 1130 | 1603 | 2551 | 3931 |
| geometry_dose | 117 | 192 | 276 | 289 | 472 |
| dose_image | 1141 | 910 | 740 | 2310 | 8332 |
| geometry_image | 2529 | 1831 | 1660 | 4694 | 8064 |
| text_geometry_dose | 82 | 168 | 260 | 240 | 382 |
| text_dose_image | 811 | 1336 | 1665 | 1286 | 3034 |
| text_geometry_image | 830 | 992 | 1274 | 1361 | 3914 |
| geometry_dose_image | 503 | 1510 | 2215 | 4148 | 4673 |
| text_dose_geometry_image | 366 | 1000 | 1067 | 1529 | 3895 |

*Table B. 5 Classification report of the best performing model over the extended dataset (text+dose+image)*

| classes | f1-score | precision | recall | support |
|---|---|---|---|---|
| axilla_ctv_approved | 1 | 1 | 1 | 27 |
| axilla_ptv_approved | 0 | 0 | 0 | 1 |
| boost_structure | 0.91 | 1 | 0.83 | 6 |
| breast_ctv_approved | 1 | 1 | 1 | 260 |
| breast_ptv_approved | 0.97 | 0.98 | 0.97 | 236 |
| chestwall_ctv_approved | 1 | 1 | 1 | 42 |
| chestwall_ptv_approved | 1 | 1 | 1 | 5 |
| combined_lungs_approved | 1 | 1 | 1 | 174 |
| combined_structure | 0.73 | 0.8 | 0.67 | 6 |
| contralateral_approved | 1 | 1 | 1 | 80 |
| control_structure | 1 | 1 | 1 | 530 |
| exclude | 0.99 | 0.99 | 1 | 1152 |
| heart_approved | 1 | 1 | 1 | 166 |
| imc_ctv_approved | 1 | 1 | 1 | 45 |

| | | | | |
|---|---|---|---|---|
| imc_ptv_approved | 1 | 1 | 1 | 5 |
| left_lung_approved | 1 | 0.99 | 1 | 172 |
| right_lung_approved | 1 | 1 | 1 | 172 |
| scf_ctv_approved | 0.99 | 1 | 0.98 | 59 |
| scf_ptv_approved | 0.5 | 1 | 0.33 | 3 |
| tumourbed_ctv_approved | 0.99 | 0.99 | 0.99 | 107 |
| tumourbed_ptv_approved | 1 | 1 | 1 | 7 |
| micro avg | 0.99 | 0.99 | 0.99 | 3255 |
| macro avg | 0.91 | 0.94 | 0.89 | 3255 |
| weighted avg | 0.99 | 0.99 | 0.99 | 3255 |
